# Fear of relapse and quality of life in multiple sclerosis: the mediating role of psychological resilience

**DOI:** 10.1101/2022.08.13.22278746

**Authors:** Yunier Broche-Pérez, Rodneys M. Jiménez-Morales, Laura Ortiz Monasterio-Ramos, Lázaro A. Vázquez-Gómez, Johana Bauer, Zoylen Fernández-Fleites

## Abstract

The goal of this study was to examine the mediating role of psychological resilience in the relationship between fear of relapse and quality of life in a sample of patients with multiple sclerosis (PwMS). This cross-sectional study was developed online. A total of 240 PwMS were surveyed using the Multiple Sclerosis Quality of Life inventory, the Fear of Relapse Scale and the Connor-Davidson Resilience Scale. To perform the mediation analysis PROCESS macro was used. In our study, fear of relapse was a predictor of psychological resilience and quality of life, and psychological resilience was a predictor of quality of life. Finally, psychological resilience showed a mediating role in the relationship between fear of relapse and quality of life. Considering that resilience is a modifiable variable, the implementation of interventions aimed at enhancing resilience can have a favorable impact on the psychological well-being and quality of life of patients with multiple sclerosis.

## Introduction

Multiple sclerosis (MS) is a chronic and demyelinating disease that significantly affects the quality of life (QoL) of people who suffer from it (Berrigan et al., 2016). A wide range of factors negatively affect the quality of life of patients with multiple sclerosis (PwMS) including fatigue, stiffness, walking difficulties, spasms, cognitive problems, sexual disorders, pain, urinary dysfunction, dizziness, emotional or mood disorders, vision problems, tremor, among others (Amato et al., 2001; Ochoa-Morales et al., 2019; Yalachkov et al., 2019).

In addition, in patients who experience relapses, a significant worsening of the quality of life has also been verified when compared with periods where there is no disease activity and also in comparison with patients who do not experience relapses at the time of evaluation (Ozakbas et al., 2004). Recently, it has also been suggested that not only does experiencing a relapse have a negative impact on the quality of life in PwMS, but the expectation of a relapse itself is also a factor that undermines the perceived quality of life in PwMS. In this sense, patients with multiple sclerosis who experience greater fear of relapse also show a poorer quality of life (Khatibi et al., 2021).

On the other hand, there are protective factors that could reduce the negative impact of fear of relapse on quality of life in patients with multiple sclerosis, such as psychological resilience. Psychological resilience is a process in which individuals display positive adjustment, despite being faced with significant adversity (Luthar & Cicchetti, 2000). In the case of MS, there is evidence that suggests that psychological resilience is linked, directly and indirectly, with multiple variables related to quality of life, such as self-efficacy, self-compassion, disability-specific variables (i.e. fatigue and physical independence), social support, positive affect and negative affect, among others (Black & Dorstyn, 2015; Nery-Hurwit et al., 2018).

However, to date we do not know of any published studies that explore the mediating effect that psychological resilience has on the relationship between fear of relapse and quality of life in PwMS. In part, the lack of studies in this field is due to the fact that until very recently there were no instruments that assessed fear of relapse in PwMS. However, a questionnaire that measures fear of relapses and disease activity in MS patients was recently developed (Khatibi et al., 2020). Since the publication of this scale, it has been proven that fear of relapse in PwMS is related to higher levels of anxiety, depression and stress, as well as a lower overall quality of life in these patients (Khatibi et al., 2021; Shaygannejad et al., 2021).

If it is found that resilience acts as a positive mediator between fear of relapse and quality of life, the result could have psychotherapeutic implications to enhance psychological well-being in PwMS. Considering that resilience is modifiable, the development of resilience-enhancing interventions could have a significant impact on the quality of life of patients with multiple sclerosis and also of other neurodegenerative diseases (Ovaska-Stafford et al., 2021).

Therefore, the main purpose of this study is to examine the mediating role of psychological resilience in the relationship between fear of relapse and quality of life in a sample of PwMS. To this end, the following hypotheses were tested:

H1. Fear of relapse will be a negative predictor of psychological resilience in the PwMS.

H2. Psychological resilience will be a positive predictor of quality of life in the PwMS.

H3. Fear of relapse will be a negative predictor of quality if life in the PwMS.

H4. Psychological resilience will have a mediating role in the relationship between fear of relapse and quality of life in the PwMS.

## Methods

### Study Design and Participants

This cross-sectional study was developed online and is part of the project «Positive modulatory variables of perceived quality of life in patients with multiple sclerosis (ME-POSITIVE project)», coordinated by the department of psychology of the Universidad Central «Marta Abreu» de Las Villas, Cuba. The survey was disseminated using Google Forms^®^. Potential participants were invited to participate in the study through social networks (Instagram, WhatsApp and Facebook groups) and direct telephone calls (using the personal information registered in the medical records). Additionally, the presidents of several patient associations were contacted, to whom the purpose of the study was explained and the survey link was shared with them. As an additional resource, one of the researchers (RMJM) participated in dissemination actions in mass media and social networks (radio programs and dissemination on Instagram) where he explained the objective of the research and patients with multiple sclerosis were invited to participate.

The study was conducted between August 13 and November 13, 2021. In total, 240 patients with multiple sclerosis completed the survey, all over 18 years of age. The study included patients from 12 Spanish-speaking countries (Argentina, Guatemala, Mexico, Dominican Republic, Chile, Spain, Cuba, Colombia, Uruguay, Paraguay, Peru, and El Salvador).

### Measures

Demographic and Clinical Information: The demographic variables explored included the age, disease duration, gender, education, marital status, country and MS phenotype.

Multiple Sclerosis Quality of Life 29 items version (MSQOL-29) (Rosato et al., 2019): The MSQOL-29 is a reduced version of the original 54-item inventory (MSQOL-54). MSQOL-29 consists of seven multi-item subscales: ‘physical function’ (six items); ‘sexual function’ (four items); ‘bodily pain’, ‘emotional well-being’, ‘energy’, ‘cognitive function’ and ‘health distress’ (three items each one); and four single-item subscales (‘social function’, ‘health perceptions’, ‘overall quality of life’, and ‘change in health’). The scale also has two composite scores (Physical Health Composite, (PHC) and Mental Health Composite (MHC)).

The Fear of Relapse Scale (Khatibi et al., 2020): The Fear of Relapse Scale is a 26-item scale grouped into three factors. Each item scored on a five-Likert scale ranging from 1 = never, 2 = rarely, 3 = sometimes, 4 = often, and 5 = always. The first factor (Fear of disability following a relapse) is composed of 13 items (items 1,2,3, 6, 7, 8, 9, 10, 11, 17, 24, 25 and 26), the second factor (Fear of the psychological and physiological consequences of a relapse) is composed of 8 items (12, 13, 14, 15, 16, 18, 20, 21) and the third factor (Limitation resulting from fear) groups items 4, 5, 19, 22 and 23 (Khatibi et al., 2020).

The Connor-Davidson Resilience Scale 10 items version (CD-RISC-10) (Campbell-Sills & Stein, 2007). The 10-item CD-RISC was extracted from the original 25-item CD-RISC (Connor & Davidson, 2003). Patients’ rate items on a 5-point Likert scale, ranging from 0 (not true at all) to 4 (true nearly all the time). Total scores for the CD-RISC-10 rage from a minimum of 0 to a maximum of 40. Total scores are calculated by summing all 10 items. A higher score indicates higher resilience.

### Ethical statement and data analysis

The study protocol was approved by the ethics committee of the Department of Psychology of the Universidad Central “Marta Abreu” de Las Villas. All procedures performed in this study were in accordance with the ethical standards of the 1964 Helsinki Declaration. Informed consent was obtained from all participants included in the study. The data were analyzed using IBM-SPSS 21.

To perform the mediation analysis, PROCESS macro was used (Hayes, 2018). We used bootstrapping sampling (n =5000) distributions to calculate the direct and indirect effects and confidence intervals (95%) of the estimated effects. Significance was determined when the confidence interval does not include zero. Before performing the mediation analysis, lack of multicollinearity, multivariate normality, and linearity were checked. The data was also checked for outliers. Normality was checked using skewness, and kurtosis values.

## Results

### Characteristics of the sample

Demographic and clinical information for 240 PwMS is summarized in Table 1. The mean age of patients (n = 240) was 40.62 years (SD ± 11.15), with a range between 18 years to 72 years old. Of the 240 MS patients 74.6% (*n* = 179) had relapsing remitting MS (RRMS) and the mean duration since disease onset was 6.86 years. In the sample, 192 participants (80.0%) were females, and 48 males (20.0%). In our sample, most of the participants had a university-level degree (55.8%). Additionally, the 42.1% of the patients were married. In the sample, patients from Argentina (27.5%), Mexico (22.9%) and Uruguay (13.8%) predominated.

**Table 1.** Participant demographic and clinical data (n = 240)

| Demographic variable | Mean ±SD or n(%) |
| --- | --- |
| Age | 40.62±11.15 |
| Disease duration years | 6.86±7.51 |
| MS Phenotype |  |
| Relapsing-Remitting (RRMS) | 179(74.6) |
| Primary-Progressive (PPMS) | 17(7.1) |
| Secondary-Progressive (SPMS) | 14(5.8) |
| Progressive-Relapsing (PRMS) | 4(1.7) |
| I Don't Know | 26(10.8) |
| Gender |  |
| Female | 192(80.0) |
| Male | 48(20.0) |
| Education |  |
| Primary education | 6(2.5) |
| Middle School | 29(12.1) |
| High School | 71(29.6) |
| University Degree | 134(55.8) |
| Marital status |  |
| Single | 92(38.3) |
| Married | 101(42.1) |
| Divorced | 45(18.8) |
| Widowed | 2(.8) |
| Country |  |
| Argentina | 66(27.5) |
| Mexico | 55(22.9) |
| Uruguay | 33(13.8) |
| Spain | 24(10.0) |
| Dominican Republic | 23(9.6) |
| Cuba | 17(7.1) |
| Colombia | 10(4.2) |
| Chile | 4(1.7) |
| Paraguay | 2(.8) |
| Guatemala | 1(.4) |
| Peru | 1(.4) |
| El Salvador | 1(.4) |

Note.SE(Standard Deviation)

### Correlation between variables

The results indicated that the skewness ranged from -.67 to .22 and kurtosis ranged from - .44 to .51 and were within the normality criteria (Byrne, 2016; Hair et al., 2010). A Durbin-Watson test r was 1.9, indicating no autocorrelation (Field, 2013). Our results showed an inverse correlation between fear of relapse and QoL (r = -.44, p <.001), indicating that increasing fear of relapse decreases the perceived quality of life in PwMS. We also found a strong (positive) correlation between psychological resilience and quality of life, with high levels of resilience associated with high levels of QoL among our sample (*r* = .32, *p* < .001). An inverse correlation was also found between fear of relapse and psychological resilience, verifying that as fear of relapse increased, resilience decreased and vice versa (r = -.23, p <.001) (table 2).

**Table 2.**
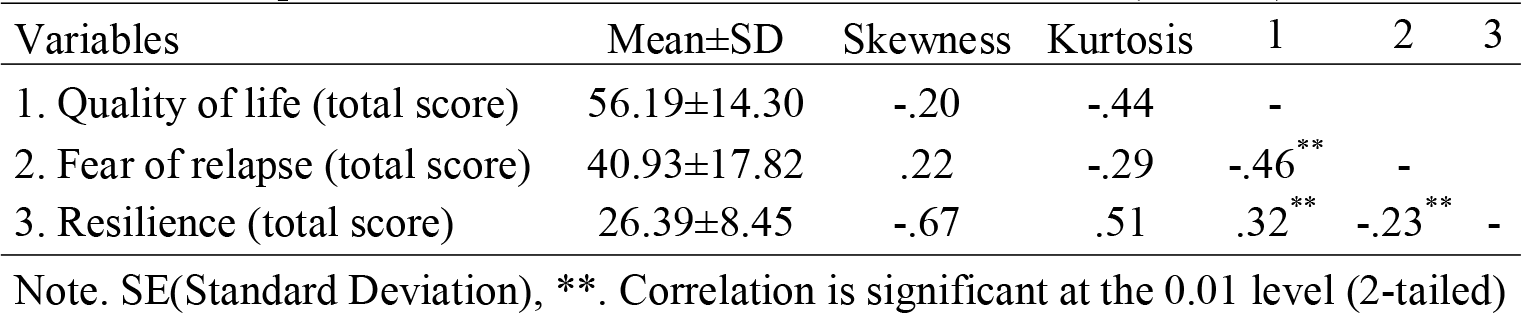
Descriptive statistics and correlation between variables (n = 240)

### Mediation analysis

The direct effect from fear of relapse to psychological resilience (path a) was significant (*β* = -.10, 95% CI [-.16, -.04] t = -3.57, p = .0004), supporting hypothesis 1. Increasing levels of fear of relapse is related to a decrease in resilience levels in our PwMS. Additionally, the hypothesis 2 is also supported. The direct effect from resilience to quality of life (path b) was also significant (*β* = .39, 95% CI [.19, .58] t = 4.02, p = .003). When the levels of resilience are increased, we can also predict an increase in the QoL in our patients. On the other hand, path c’ indicates that fear of relapse predicts QoL (*β* = -.32, 95% CI [-.41, -.23] t = 7.10, p < .001), supporting hypothesis 3. When the levels of fear of relapses increase, a decrease in quality of life levels can be predicted. The total effect of fear of relapse on quality of life was significant (*β* = -.36, 95% CI [-.45, -.27] t = 7.97, p < .001), and when psychological resilience, which is the mediation variable, is included in the model, this effect (*β* =.-.36) decreases (*β* = -.32). The indirect effect of fear of relapse on QoL does not include zero *β* =-.041, 95% CI (-.084, -.010). According to this result, psychological resilience acts as a mediator between fear of relapse and quality of life in our sample of PwMS, supporting hypothesis 4 (figure 1).

**Fig. 1.**
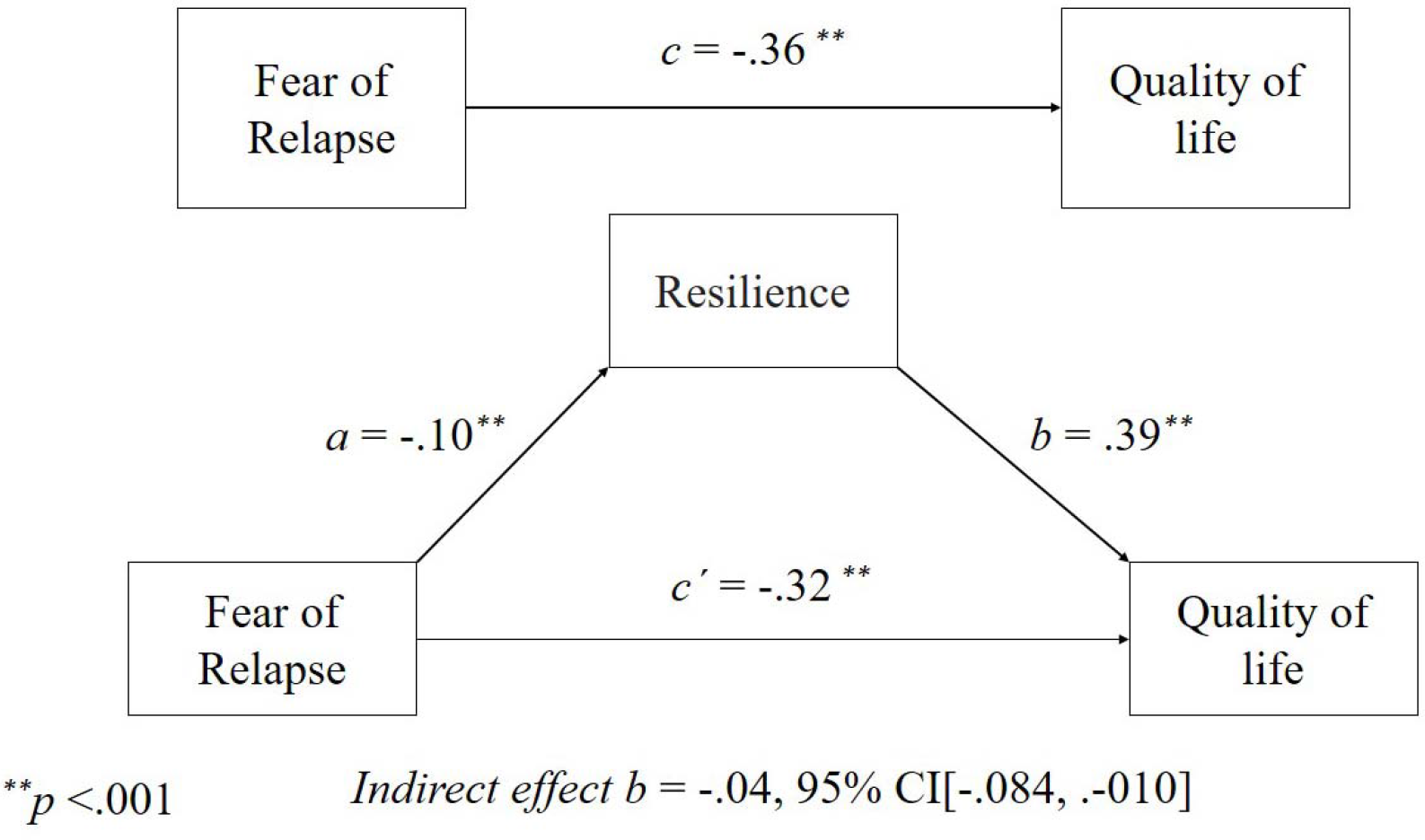
The statistical diagram of basic mediation model

## Discussion

The objective of this study was to examine the mediating role of psychological resilience in the relationship between fear of relapse and quality of life in a sample of PwMS. First, our results confirmed that, in our sample, fear of relapse constitutes a negative predictor of psychological resilience (hypothesis 1) and quality of life (hypothesis 3) in the PwMS.

The negative impact of fear of relapse on QoL of patients with multiple sclerosis has recently been explored. In a study that included 168 patients with Relapsing-Remitting MS (RRMS) it was found that PwMS who showed higher levels of fear of relapse also reported higher levels of intolerance of uncertainty, depression, anxiety and stress (Khatibi et al., 2020). According to the study authors, these results would be related to a greater fear of disability that would lead to a new relapse. In this sense, there is consensus that a new relapse is a sign of disease activity and has its implications such as treatment failure or accumulation of disability (Khatibi et al., 2020), an expectation that would negatively impact the physical and psychological well-being of PwMS.

It has also been reported that patients with multiple sclerosis who report higher levels of fear of relapse experience more anxiety related to health status and a lower QoL in the dimension related to physical health. These were the conclusions reached by a study that explored the relationship between fear of relapse and quality of life in a sample of 70 RRMS patients (Khatibi et al., 2021).

On the other hand, our results confirm that psychological resilience is a positive predictor of quality of life in the PwMS (hypothesis 2). Our results support the conclusions of a study conducted by Ploughman et al. (2020), in a sample of 743 patients with multiple sclerosis. In this study, the patients were divided into two groups, a group with high levels of resilience and another with low levels. Patients with high levels of resilience lived with less disability and fatigue, reported greater participation, exercised more, consumed a healthier diet and lived with greater social support and financial security, compared to the lower scoring group (Ploughman et al., 2020).

It has also been suggested that high psychological resilience is associate with better objective performance on the MS Functional Composite (MSFC) and motor outcomes, especially gross motor function (i.e., grip strength, gait endurance) (Klineova et al., 2020). To explore psychological resilience in these patients, the authors used the same scale used by us in the present study (CD-RISC-10), verifying that this relationship of psychological resilience to outcomes was independent of mood and fatigue, and psychological resilience was unrelated to disease burden or traditional metrics of neurologic disturbance (for example, EDSS).

Our study also revealed the significant mediating role of resilience in the relationship between fear of relapses and QoL in PwMS (hypothesis 4). The mediating nature of resilience on the quality of life of PwMS has been previously reported by other authors. For example, in a study conducted by Nery-Hurwit et al. (2018) in a sample of 259 PwMS, the roles of self-compassion and resilience on perceived health-related quality of life for individuals with multiple sclerosis were explored using mediation analysis. The conclusions of the study showed a significant direct effect between self-compassion and health-related quality of life as well as an indirect relationship through resilience. According to the authors increasing resilience may help individuals overcome stressful and traumatic events and experience quality of life with disability.

When other variables such as age, sex, psychological health and severity of disease symptoms are controlled for, resilience has also shown a positive association with quality of life in people with MS (Battalio et al., 2017). Similarly, it has been found that in persons with long-term disabilities (including PwMS) who have high levels of resilience, there is a lower impact of secondary symptoms such as pain and fatigue on quality of life (Terrill et al., 2016).

Recently, Kasser and Zia (2020) examined the relationship between disease-related risk factors, protective factors coping and resilience on QoL in adults with MS (n = 271), using a structural equation modeling approach. Disability level, fatigue, walking impairment, fear of falling, falls, and pain were considered as risk factors. On the other hand, physical activity, self-efficacy, social support, optimism, and health locus of control were modeled as protective factors. Psychological resilience and coping were modeled as two latent variables. The results of the study showed that while risk factors decreased quality of life, resilience increased the likelihood of higher quality of life in the sample of PwMS (Kasser & Zia, 2020).

In general, these evidences show that psychological resilience modulates the quality of life in PwMS. In the particular case of fear of relapse, psychological resilience could be interacting with other variables such as controllability, reducing anxiety related to anticipating a new relapse of the disease. Furthermore, there is evidence that external factors of resilience, such as satisfaction with social roles, social support, social functioning, and social connection, also positively modulate quality of life in people with neurodegenerative diseases, including PwMS (Ovaska-Stafford et al., 2021).

This study is not without its limitations. First, this is a cross-sectional study so it is difficult to accurately elucidate causal relationships between fear of relapse, psychological and quality of life. Another limitation of the present study is the heterogeneity of the phenotypes included in the sample. In this sense, the levels of fear of relapse could vary considerably between clinical phenotypes. For example, patients with Relapsing-Remitting MS (RRMS) may experience less fear of relapses, a better overall quality of life compared to patients with progressive forms of the disease, which could also influence resilience and quality of life. Thus, future studies should explore the relationships between the variables studied taking into account the clinical characteristics of each phenotype. Finally, we consider that an important limitation of the research is that it does not take into account other variables potentially modulating the quality of life in PwMS, such as the level of fatigue and cognitive functioning. This is because the study design explores a simple mediation relationship, for which reason other relevant variables should be included in future studies.

Despite these limitations, our results offer evidence on the importance of resilience as a modulating factor of quality of life in patients with multiple sclerosis. Considering that resilience is a modifiable variable, the implementation of interventions aimed at enhancing resilience can have a favorable impact on the psychological well-being of patients with multiple sclerosis.

## Data Availability

All data produced in the present study are available upon reasonable request to the authors.

## Statements and Declarations

### Ethical standards

All procedures followed were in accordance with the ethical standards of the responsible committee on human experimentation (institutional and national) and with Helsinki Declaration of 1975, as revised in 2000. Informed consent was obtained for all patients for being included in the study.

### Declaration of Competing Interest

The authors have no conflicts of interest

### Funding

This research did not receive any specific grant from funding agencies in the public, commercial, or not-for-profit sectors.

## Acknowledgement

First of all we want to express our gratitude to all the people with MS who participated in the study. We especially thank the coordinators and patients of the EMA (Esclerosis Múltiple Argentina) and ALCEM (Asociación de Lucha Contra la Esclerosis Múltiple) associations, as well as the patient associations of Mexico, Uruguay, Cuba and the Dominican Republic. We also thank Rocio Seijas (Escleroamigos) and Fernando Champomier (Emstrongs) who from their projects supported our research from the beginning. Finally, we especially thank Nicolás Edgardo Costa, coordinator of the program «The game does not end» (ALCEM podcast series made by people living with MS).

## Notes

### Competing Interest Statement

The authors have declared no competing interest.

### Funding Statement

This study did not receive any funding.

### Author Declarations

The study protocol was approved by the ethics committee of the Department of Psychology of the Universidad Central Marta Abreu de Las Villas. All procedures performed in this study were in accordance with the ethical standards of the 1964 Helsinki Declaration. Informed consent was obtained from all participants included in the study.

## Referencias

Amato, M. P., Ponziani, G., Rossi, F., Liedl, C. L., Stefanile, C., & Rossi, L. (2001, Oct). Quality of life in multiple sclerosis: the impact of depression, fatigue and disability. Mult Scler, 7(5), 340–344. https://doi.org/10.1177/135245850100700511

Battalio, S. L., Silverman, A. M., Ehde, D. M., Amtmann, D., Edwards, K. A., & Jensen, M. P. (2017, Jun). Resilience and Function in Adults With Physical Disabilities: An Observational Study. Arch Phys Med Rehabil, 98(6), 1158–1164. https://doi.org/10.1016/j.apmr.2016.11.012

Berrigan, L. I., Fisk, J. D., Patten, S. B., Tremlett, H., Wolfson, C., Warren, S., Fiest, K. M., McKay, K. A., & Marrie, R. A. (2016, Apr 12). Health-related quality of life in multiple sclerosis: Direct and indirect effects of comorbidity. Neurology, 86(15), 1417–1424. https://doi.org/10.1212/wnl.0000000000002564

Black, R., & Dorstyn, D. (2015, Nov). A biopsychosocial model of resilience for multiple sclerosis. J Health Psychol, 20(11), 1434–1444. https://doi.org/10.1177/1359105313512879

Byrne, B. (2016). Structural Equation Modeling With AMOS: Basic Concepts, Applications, and Programming, Third Edition (3rd ed.). Routledge

Campbell-Sills, L., & Stein, M. B. (2007, Dec). Psychometric analysis and refinement of the Connor-davidson Resilience Scale (CD-RISC): Validation of a 10-item measure of resilience. J Trauma Stress, 20(6), 1019–1028. https://doi.org/10.1002/jts.20271

Connor, K. M., & Davidson, J. R. (2003). Development of a new resilience scale: the Connor-Davidson Resilience Scale (CD-RISC). Depress Anxiety, 18(2), 76–82. https://doi.org/10.1002/da.10113

Field, A. (2013). Discovering statistics using IBM SPSS statistics (4th ed.). SAGE Publications.

Hair, J., Black, W. C., Babin, B. J., & Anderson, R. E. (2010). Multivariate data analysis (Vol. 7th). Pearson Educational International.

Hayes, A. F. (2018). Introduction to mediation, moderation, and conditional process analysis (2nd ed.). The Guilford Press.

Kasser, S. L., & Zia, A. (2020, Jul). Mediating Role of Resilience on Quality of Life in Individuals With Multiple Sclerosis: A Structural Equation Modeling Approach. Arch Phys Med Rehabil, 101(7), 1152–1161. https://doi.org/10.1016/j.apmr.2020.02.010

Khatibi, A., Moradi, N., Rahbari, N., Salehi, T., & Dehghani, M. (2020, 2020-March-20). Development and Validation of Fear of Relapse Scale for Relapsing-Remitting Multiple Sclerosis: Understanding Stressors in Patients [Original Research]. Frontiers in Psychiatry, 11(226). https://doi.org/https://doi.org/10.3389/fpsyt.2020.00226

Khatibi, A., Weiland, T. J., & Dehghani, M. (2021, 2021/09/01/). Fear of relapse in patients suffering from RRMS influence their quality of life. Mult Scler Relat Disord, 54, 103137. https://doi.org/https://doi.org/10.1016/j.msard.2021.103137

Klineova, S., Brandstadter, R., Fabian, M. T., Sand, I. K., Krieger, S., Leavitt, V. M., Lewis, C., Riley, C. S., Lublin, F., Miller, A. E., & Sumowski, J. F. (2020, Aug). Psychological resilience is linked to motor strength and gait endurance in early multiple sclerosis. Mult Scler, 26(9), 1111–1120. https://doi.org/10.1177/1352458519852725

Luthar, S. S., & Cicchetti, D. (2000, Autumn). The construct of resilience: implications for interventions and social policies. Dev Psychopathol, 12(4), 857–885. https://doi.org/10.1017/s0954579400004156

Nery-Hurwit, M., Yun, J., & Ebbeck, V. (2018, Apr). Examining the roles of self-compassion and resilience on health-related quality of life for individuals with Multiple Sclerosis. Disabil Health J, 11(2), 256–261. https://doi.org/10.1016/j.dhjo.2017.10.010

Ochoa-Morales, A., Hernández-Mojica, T., Paz-Rodríguez, F., Jara-Prado, A., Trujillo-De Los Santos, Z., Sánchez-Guzmán, M. A., Guerrero-Camacho, J. L., Corona-Vázquez, T., Flores, J., Camacho-Molina, A., Rivas-Alonso, V., & Dávila-Ortiz de Montellano, D. J. (2019, Nov). Quality of life in patients with multiple sclerosis and its association with depressive symptoms and physical disability. Mult Scler Relat Disord, 36, 101386. https://doi.org/10.1016/j.msard.2019.101386

Ovaska-Stafford, N., Maltby, J., & Dale, M. (2021, Feb 12). Literature Review: Psychological Resilience Factors in People with Neurodegenerative Diseases. Arch Clin Neuropsychol, 36(2), 283–306. https://doi.org/10.1093/arclin/acz063

Ozakbas, S., Cagiran, I., Ormeci, B., & Idiman, E. (2004, Mar 15). Correlations between multiple sclerosis functional composite, expanded disability status scale and health-related quality of life during and after treatment of relapses in patients with multiple sclerosis. J Neurol Sci, 218(1-2), 3–7. https://doi.org/10.1016/j.jns.2003.09.015

Ploughman, M., Downer, M. B., Pretty, R. W., Wallack, E. M., Amirkhanian, S., & Kirkland, M. C. (2020, Oct). The impact of resilience on healthy aging with multiple sclerosis. Qual Life Res, 29(10), 2769–2779. https://doi.org/10.1007/s11136-020-02521-6

Rosato, R., Testa, S., Bertolotto, A., Scavelli, F., Giovannetti, A. M., Confalonieri, P., Patti, F., Chisari, C. G., Lugaresi, A., Pietrolongo, E., Grasso, M. G., Rossi, I., Toscano, A., Loera, B., Giordano, A., & Solari, A. (2019, May). eMSQOL-29: Prospective validation of the abbreviated, electronic version of MSQOL-54. Mult Scler, 25(6), 856–866. https://doi.org/10.1177/1352458518774935

Shaygannejad, V., Mirmosayyeb, O., Nehzat, N., & Ghajarzadeh, M. (2021, Jul). Fear of relapse, social support, and psychological well-being (depression, anxiety, and stress level) of patients with multiple sclerosis (MS) during the COVID-19 pandemic stage. Neurol Sci, 42(7), 2615–2618. https://doi.org/https://doi.org/10.1007/s10072-021-05253-8

Terrill, A. L., Molton, I. R., Ehde, D. M., Amtmann, D., Bombardier, C. H., Smith, A. E., & Jensen, M. P. (2016, May). Resilience, age, and perceived symptoms in persons with long-term physical disabilities. J Health Psychol, 21(5), 640–649. https://doi.org/10.1177/1359105314532973

Yalachkov, Y., Soydaş, D., Bergmann, J., Frisch, S., Behrens, M., Foerch, C., & Gehrig, J. (2019, May). Determinants of quality of life in relapsing-remitting and progressive multiple sclerosis. Mult Scler Relat Disord, 30, 33–37. https://doi.org/10.1016/j.msard.2019.01.049

